# A survey on retracted articles in the field of health sciences from 2001 to 2022 from Iran

**DOI:** 10.1101/2023.12.07.23299438

**Authors:** Parisa Soltani, Shakiba Sadeghizade, Mohammad Hossein Nikbakht, Arghavan Malaz

## Abstract

**Background:** Retraction is the process of correcting literature and warning readers of publications that contain such serious flaws or erroneous data. A variety of reasons may lead to retraction of published articles.

**Objectives:** This study aimed to investigate the characteristics of retracted articles from Iran in the field of health sciences.

**Design:** Retrospective study

**Setting:** This study was conducted on retracted articles in the field of health published by authors affiliated with Iranian organizations from 2001 to 2022. The Retraction Watch Database was used to extract the information from these articles. The following information was extracted: journal name, article type, institutions, country(s), date published, date retracted, authors number, and reasons for retraction. This data was evaluated using descriptive statistics.

**Results:** Between 2001 and 2022, our search identified 348 retracted articles. The highest number of retractions belongs to Diagnostic Pathology and Tumor Biology journals. The highest numbers of retractions belong to 2016 and 2020, respectively. The highest number of retracted articles were published in 2015 and 2018, respectively. Different types of articles have been assigned different percentages: Research article (52.9%), clinical study (29.9%), conference abstract/paper (5.2%), review article (4.6%), case report (3.7%), and others (3.7%). The main reasons that led to retraction were investigation by journal/publisher, fake peer review, and concerns/issues about data. Malaysia, Bahrein, and India had the most cooperation with Iran in the retracted articles. The mean time between publication and retraction was 2.44 years and ranged between less than a year and 16 years. Institutions with the highest record of retraction are Kashan University of Medical Sciences, Shaheed Beheshti University of Medical Sciences, and Tabriz University of Medical Sciences.

**Conclusions:** Investigation by journal/publisher, fake peer review, and concerns/issues about data were the most common reasons for retraction. The results of this study indicate that more attention is required from research ethics authorities in order to protect the integrity of published research.

---

Strengths and Limitations: This study examines a long period of time and is entirely in the field of public health. It is possible that the site we used does not include all journals.

## Background

Retraction is the process of correcting the published literature that contains serious flaws or erroneous data. Unreliable data can be the result of honest error or research misconduct (1).

Occasionally, authors try to exploit the academic publishing system to publish articles that would otherwise not be accepted for publication. As such, they aim to achieve undeserved success for a variety of reasons, such as career advancement, greater satisfaction, and gaining credibility or respect in the scientific community, among other reasons. These days, millions of authors are trying to publish their works, and the research and success of many of them have raised expectations for scientists. What would have constituted a major achievement in terms of the number of articles published twenty years ago is now hardly considered the minimum, or even less than the minimum, for scientific achievement. Some researchers eventually succumb to social or academic pressures and choose an unethical path (2).

A published article that is later proven not worth publishing may be invalidated for certain reasons. Retraction should be issued with caution and only for appropriate cases. Committee On Publication Ethics (COPE) suggests that journal editors should consider retracting an article in one of the following situations: (a) the editors have evidence that the findings are unreliable, either as a result of misconduct or an honest mistake; (b) the same findings have already been published and this fact has not been properly addressed (e.g. by obtaining permission or providing a reference; (c) plagiarizes another published article; (d) the article is based on unethical research. According to these COPE guidelines, an article can be disqualified due to “honest errors,” “misattribution,” “data manipulation,” “poor data management” (such as the author’s inability to produce data to support his conclusions), “academic plagiarism” (including plagiarism from oneself), “duplication of text” or “non-disclosure of conflicts of interest” should be invalidated (3).

Previous articles investigated the characteristics of retracted articles from other sources such as Scopus and PubMed until 2019 (2). This study aimed to investigate the characteristics of retracted articles from Iran in the field of health sciences from 2001 to 2022.

## Setting

This study was approved by the Research Ethics Committee of Isfahan University of medical sciences (IR.MUI.RESEARCH.REC.1402.127).

This study cross-sectional was conducted on the retracted articles in the field of health sciences that were published by authors affiliated with Iranian organizations from 2001 to 2022. The Retraction Watch Database was used to extract the information of these articles (4). Iran and health sciences filters have been applied to search on this database.

The following information was extracted from the retrieved records:

- Journal name
- Article type
- Institutions
- Country(s)
- Date published
- Date retracted
- Authors number
- Reasons

The data was entered into SPSS (version 26, IBM Statistics, NY, USA) and descriptive statistics were used.

## Results

Between January 2001 and December 2022, our search identified 348 retracted articles.

Table 1 demonstrates different information about journals with the highest number of retracted articles including the number of retracted articles, CiteScore, impact factor, H-index, and subject category. The highest number of retractions belongs to Diagnostic Pathology followed by Tumor Biology.

**Table 1.**
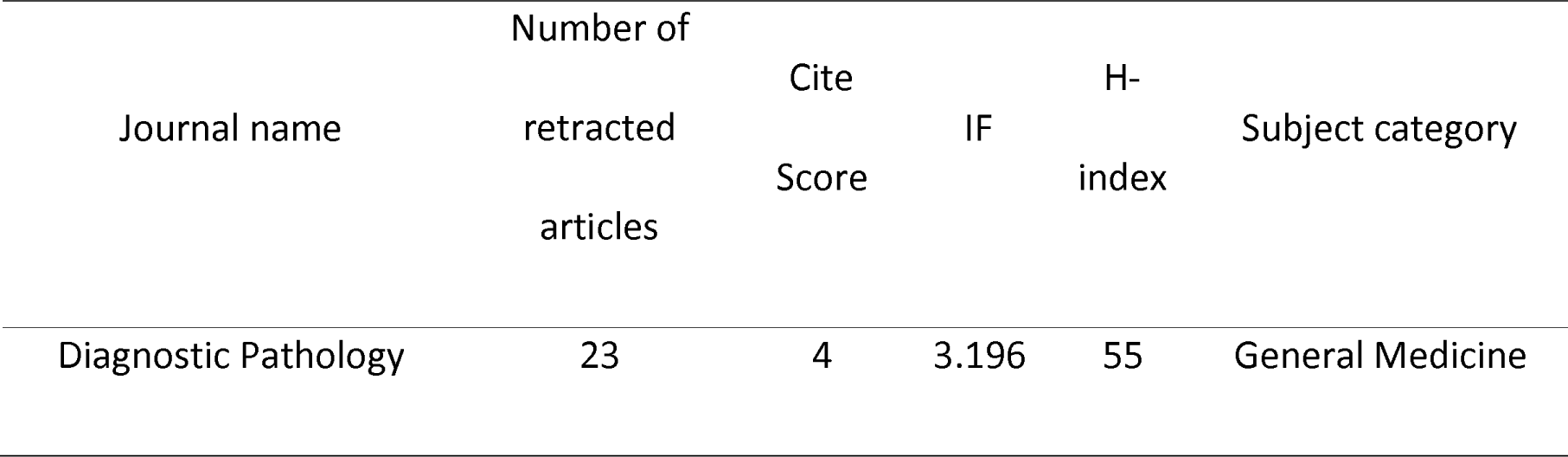
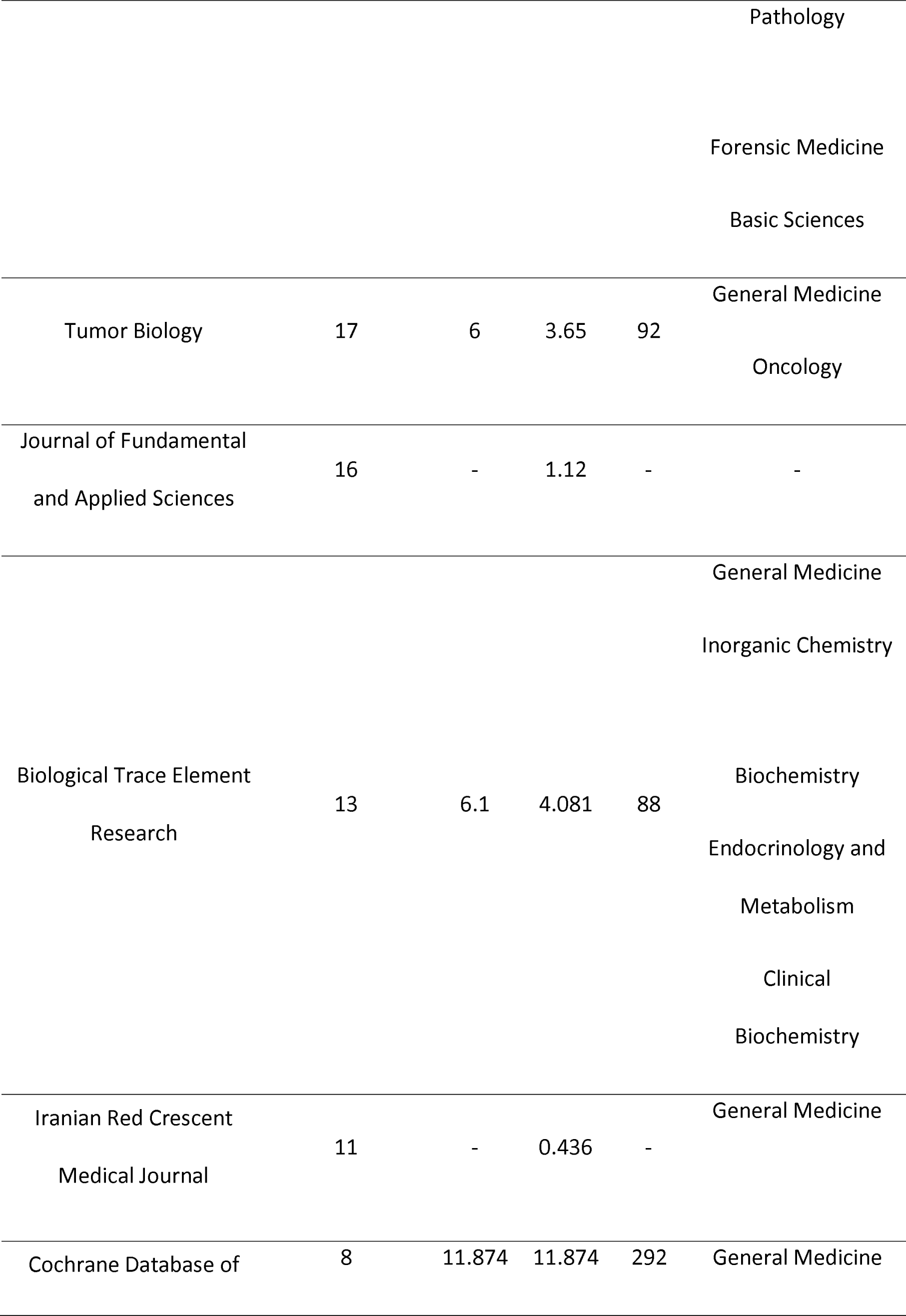
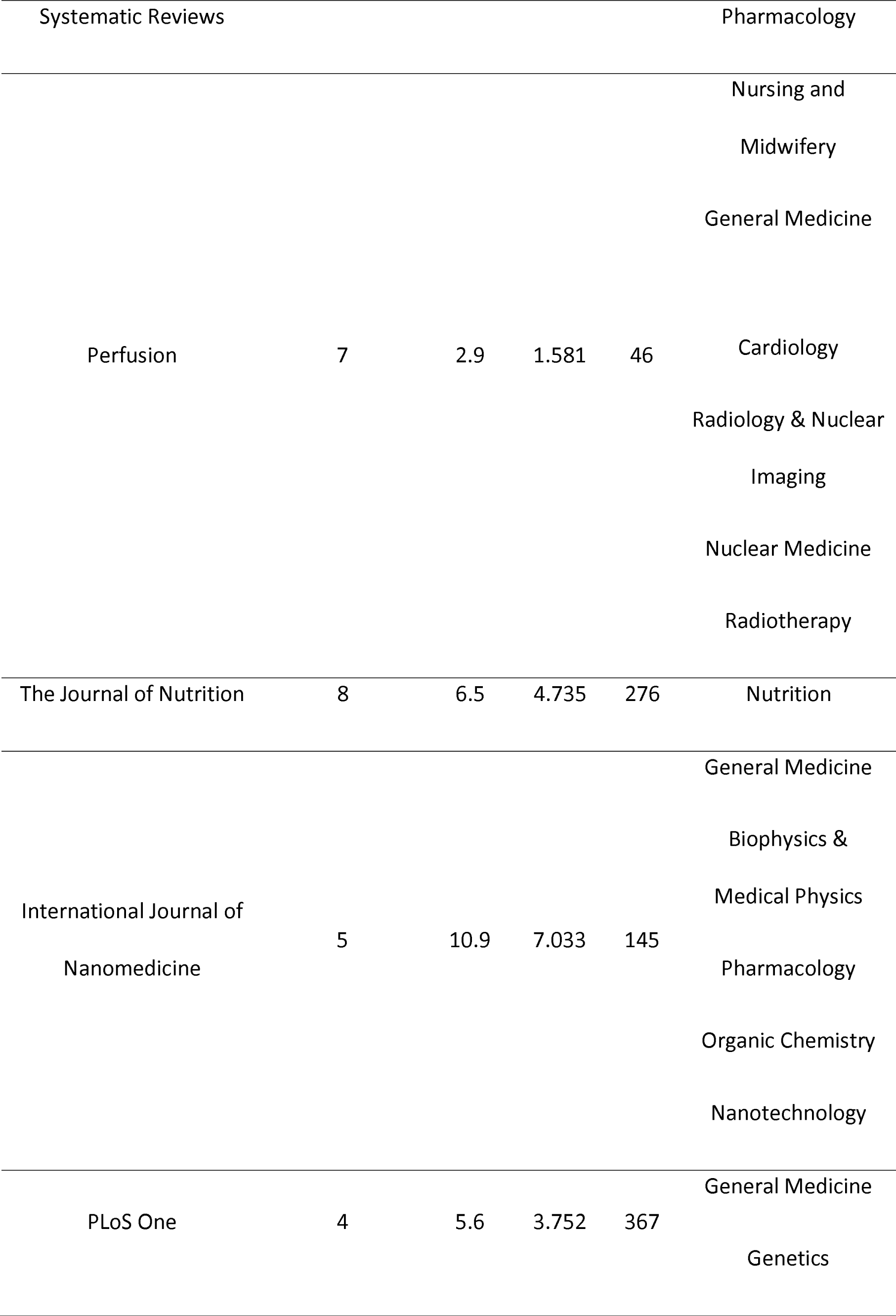
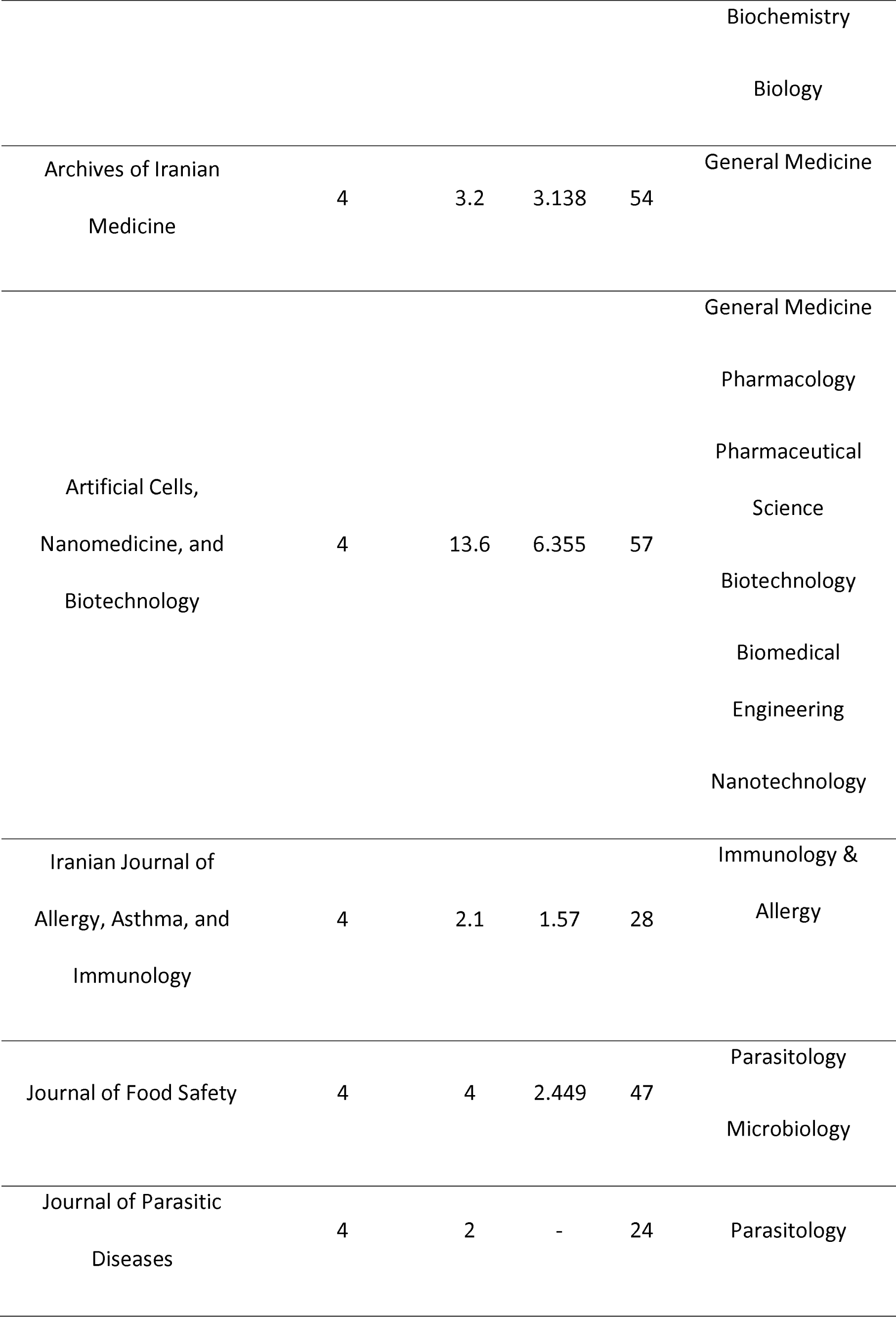
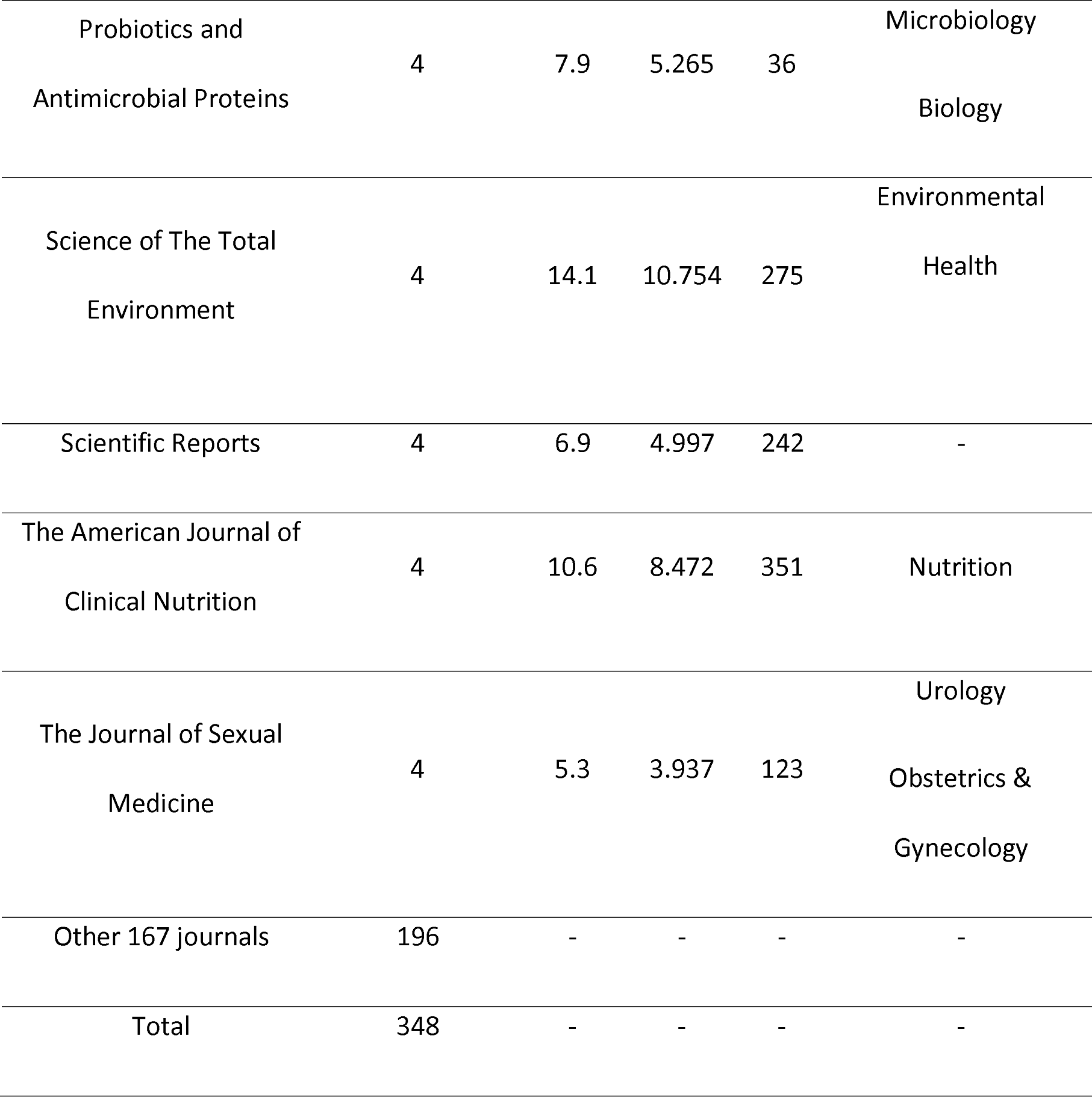
Top 20 journal information based on number of retracted articles.

Figure 1 shows the number of retracted articles based on publication year and retraction year. The highest numbers of retractions were finalized in 2016 and 2020 with 77 and 60 retractions, respectively. The most retracted articles were published in 2015 and 2018 with 58 and 48 articles, respectively.

**Figure 1.**
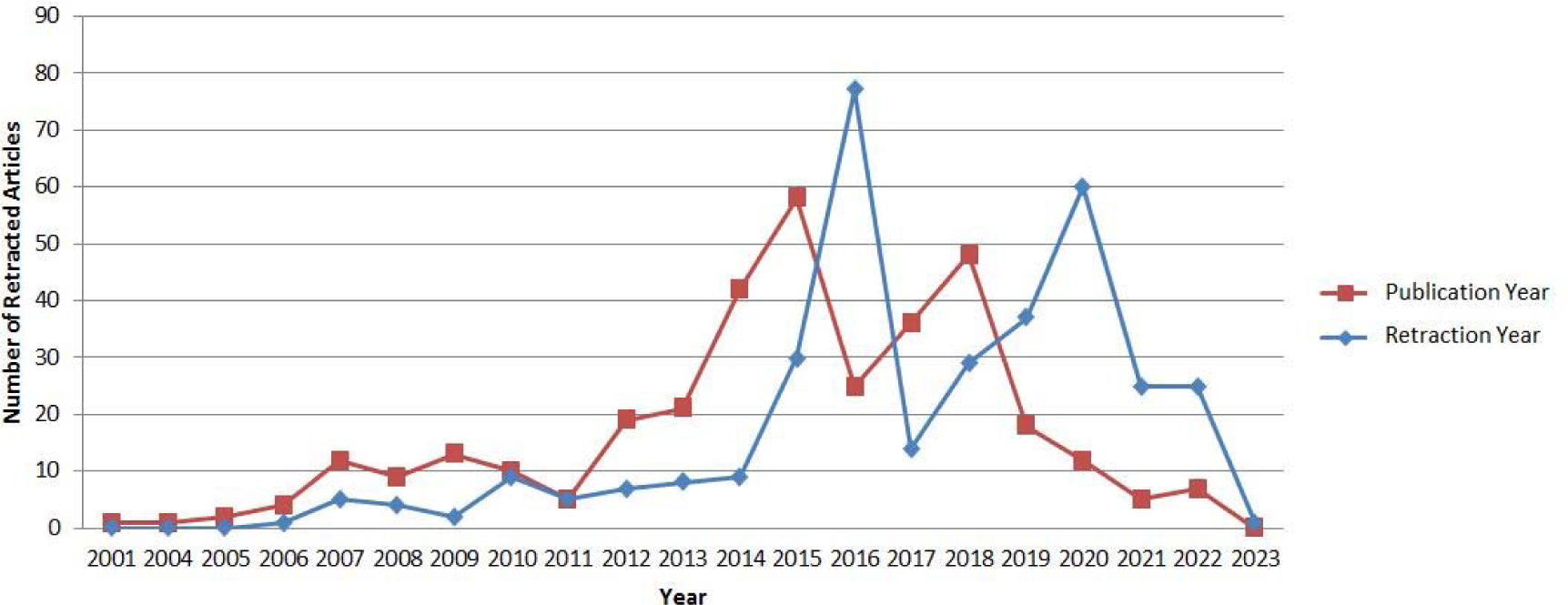
>The number of retracted articles based on publication year and retraction year.

Research articles (52.9%) and clinical studies (29.9%) were the most commonly retracted types of articles (Figure 2).

**Figure 2.**
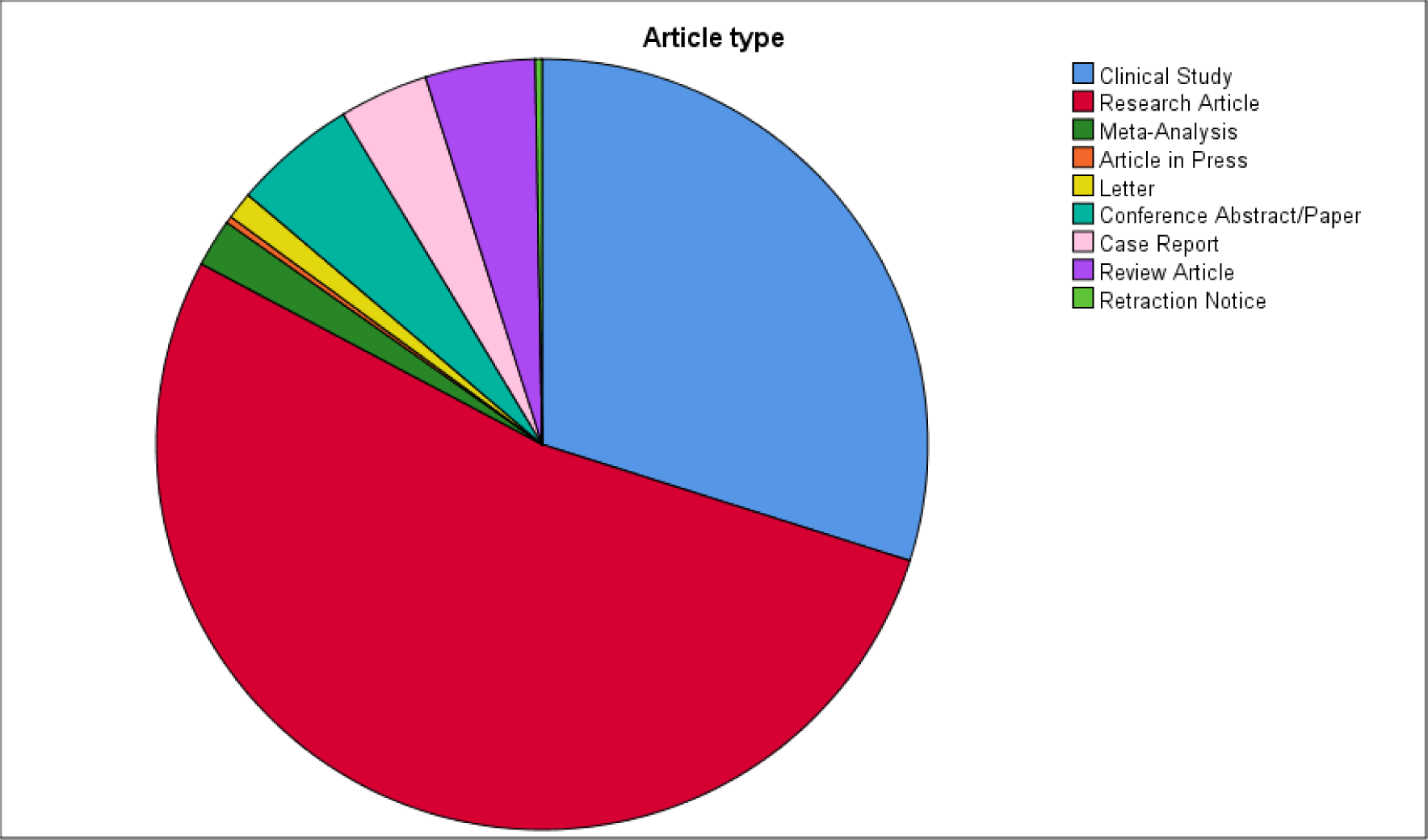
Different types of retracted articles based on quantity.

The main reasons which led to retraction are investigation by journal/publisher, fake peer review, and concerns /issues about data (Table 2).

**Table 2.**
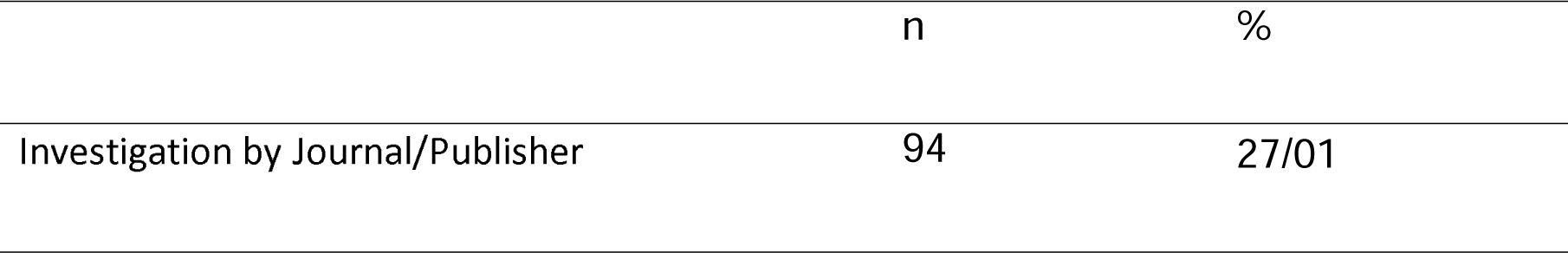
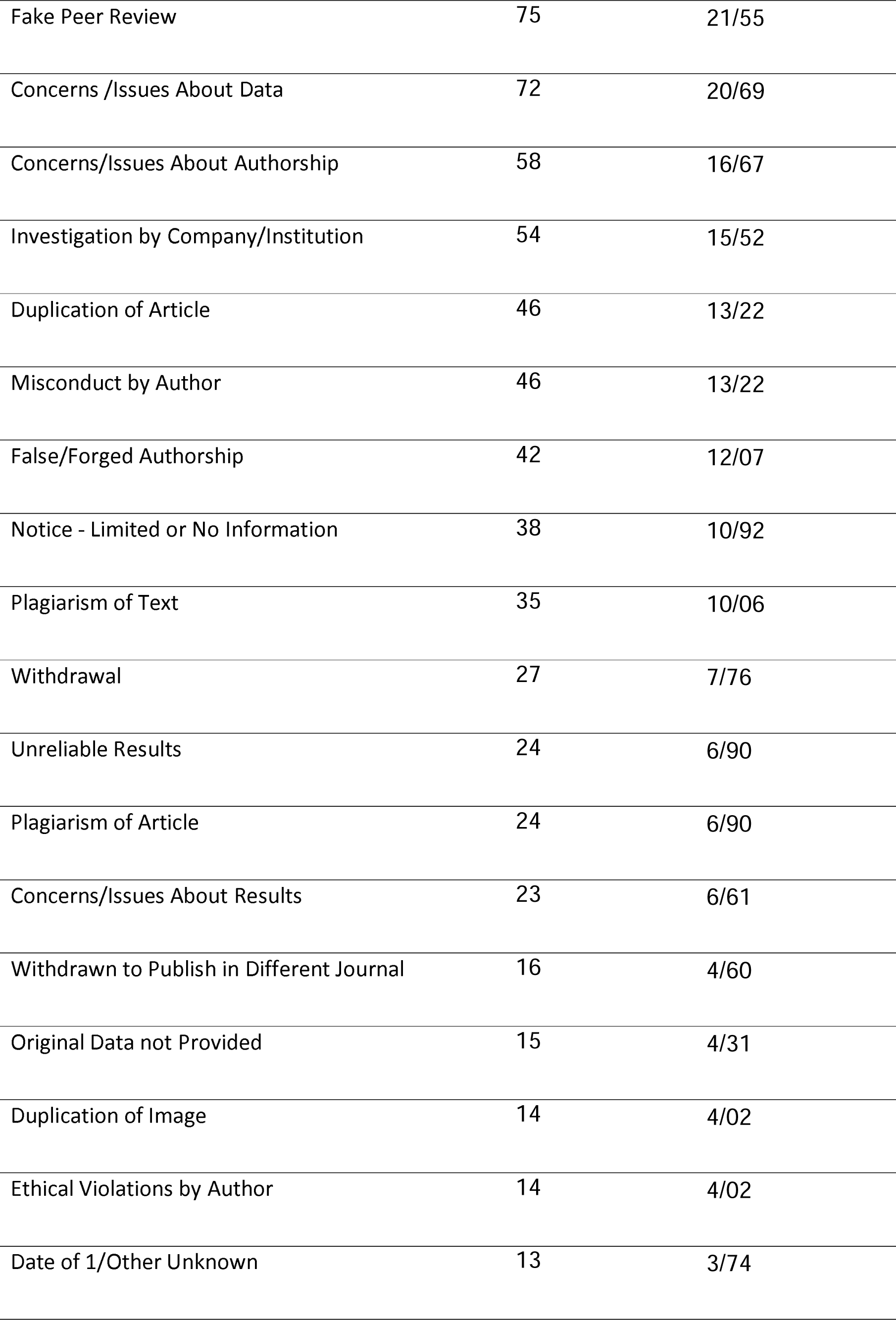
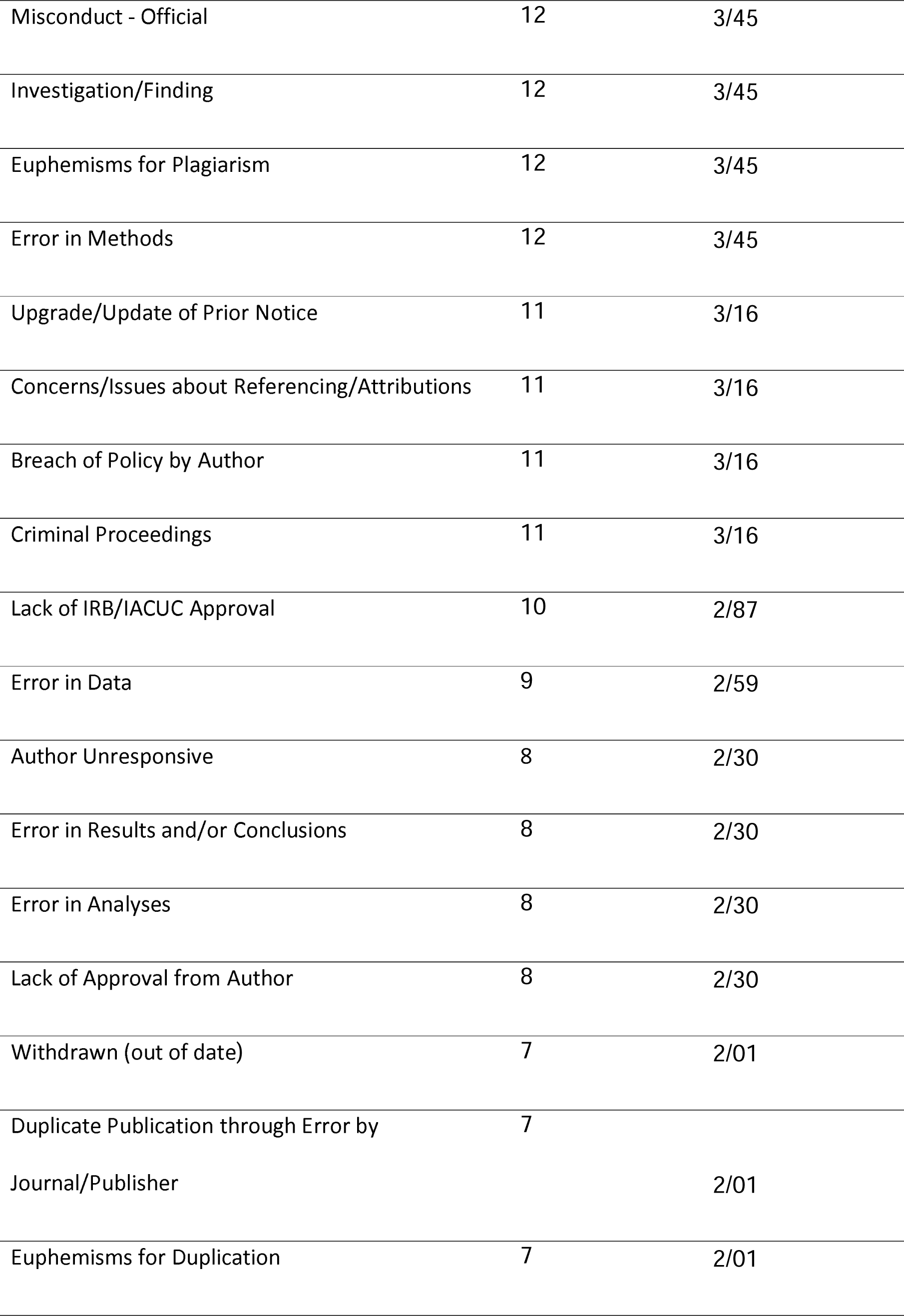
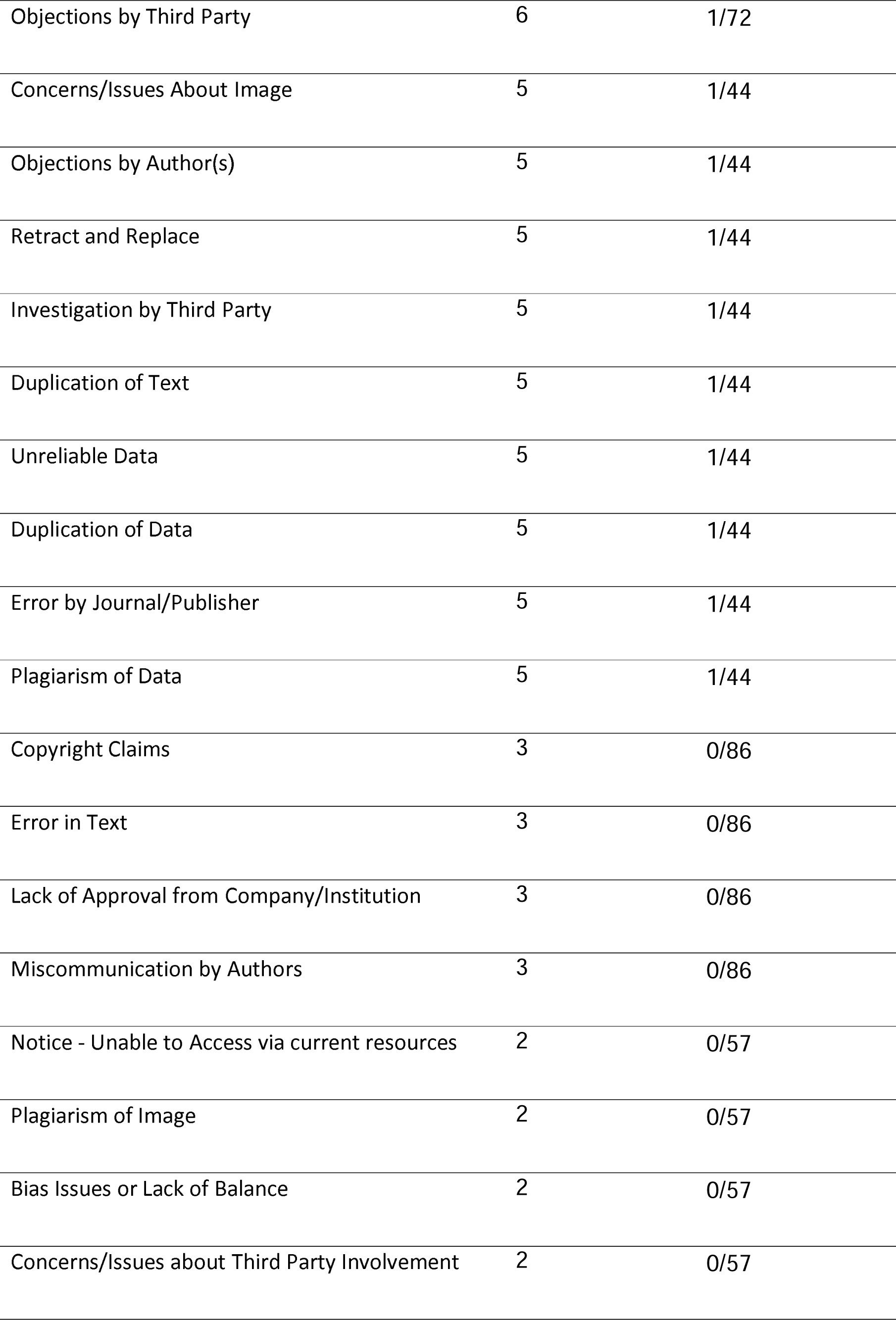
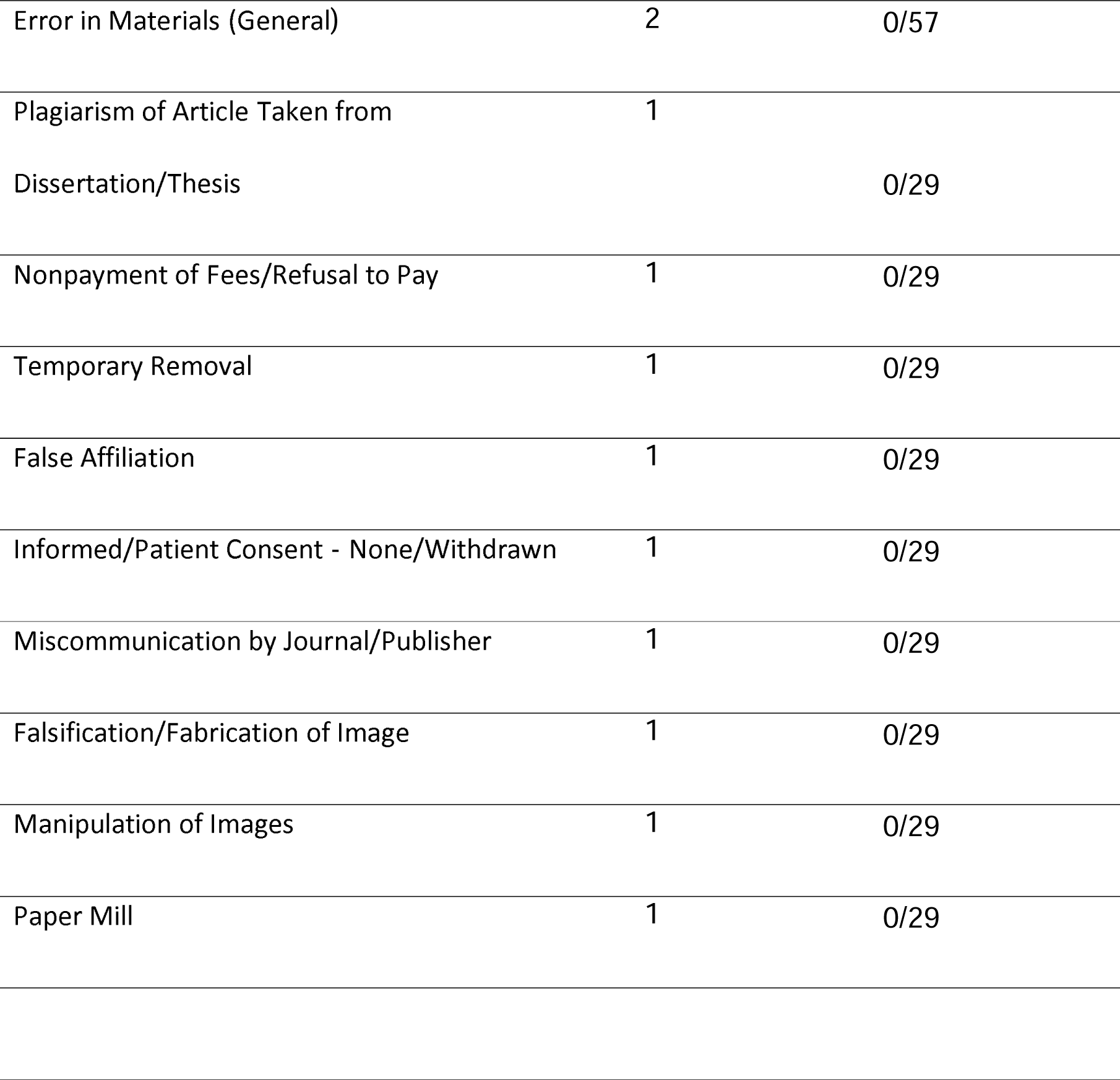
Reasons led to retraction.

Figure 3 shows the countries with most cooperation with Iran. According to the chart, Malaysia, Bahrein and India shows the highest number.

**Figure 3.**
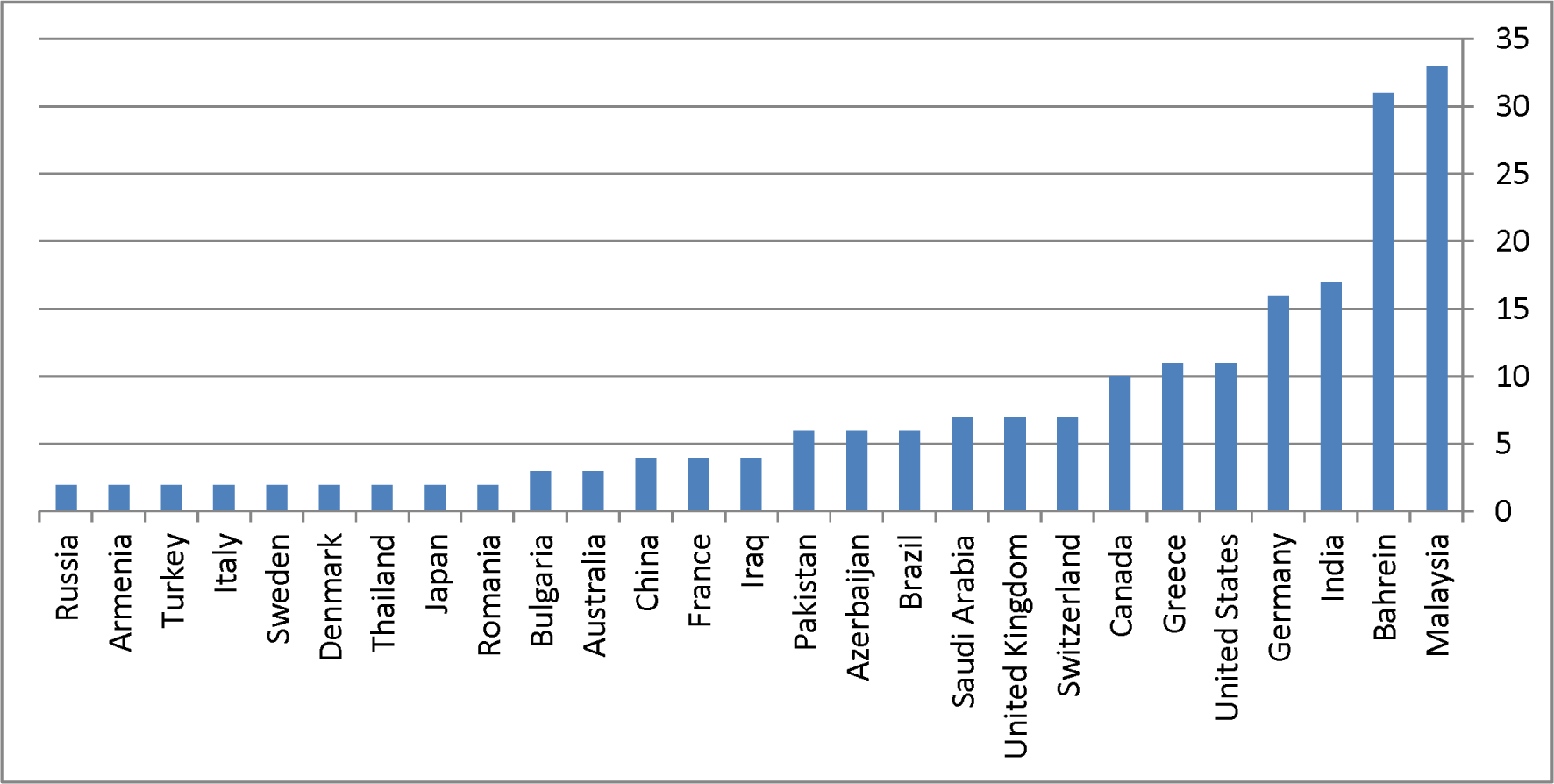
Top countries cooperated with Iran which led to retraction.

Figure 4 shows the frequency of retracted articles based on their time lag (the period of time between retraction and publication year). The mean ± SD time lag was 2.44±2.81 years and ranged between less than a year and 16 years.

**Figure 4.**
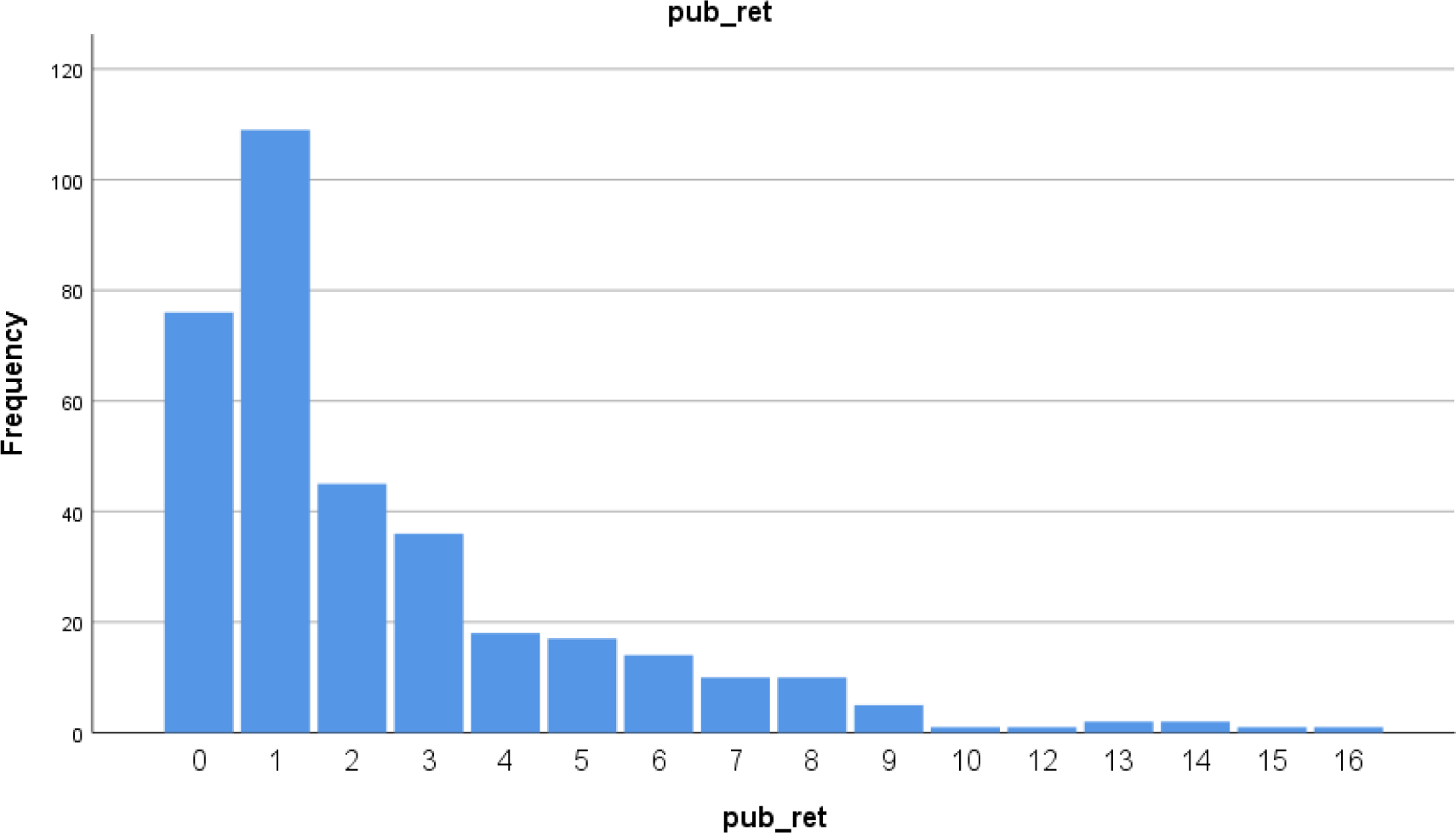
Frequency of retracted articles based on their time lag.

The institutions with the highest number of retracted papers are Kashan University of Medical Sciences, Shaheed Beheshti University of Medical Sciences, and Tabriz University of Medical Sciences (Table 3).

**Table 3.**
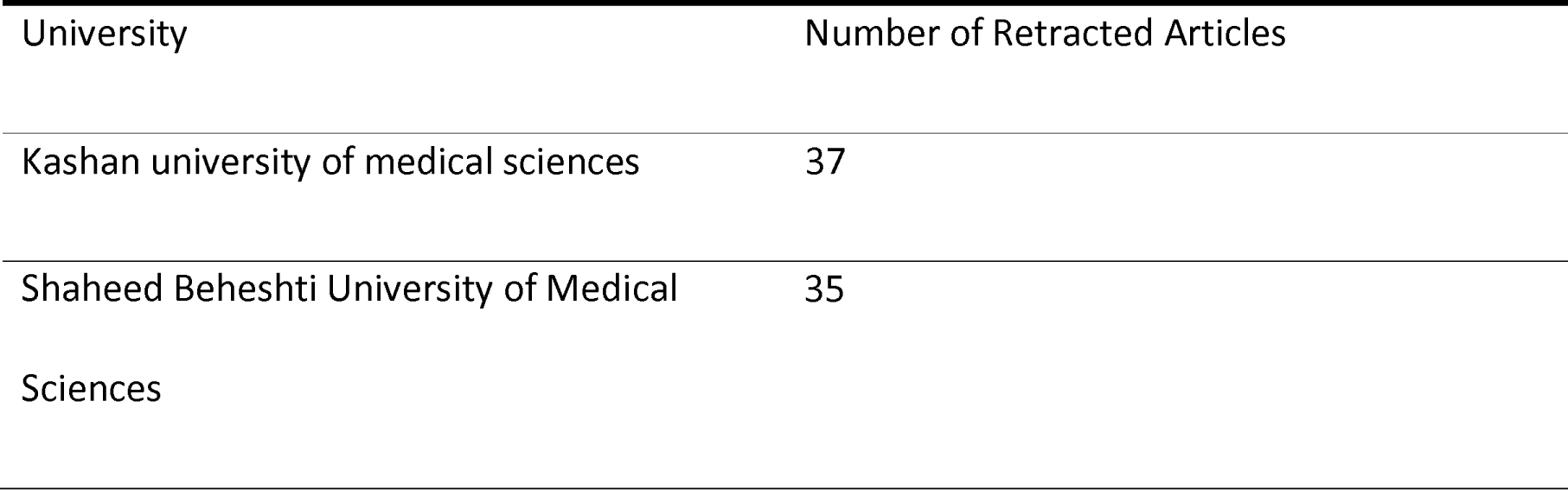
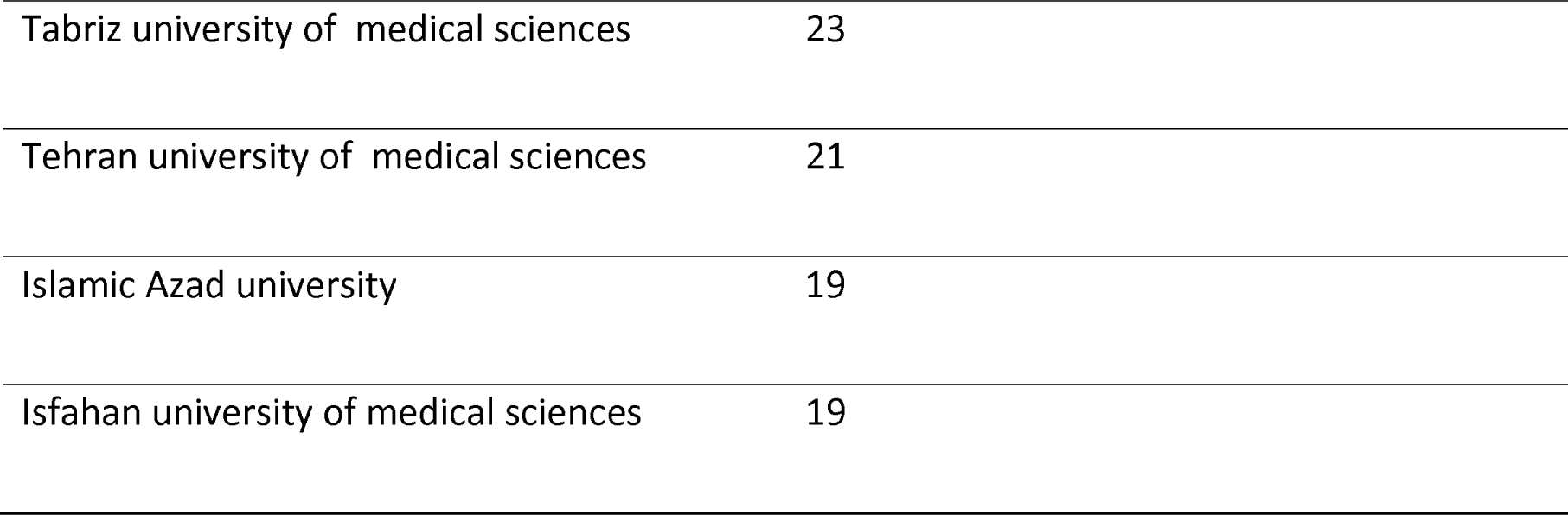
Universities with highest number of retracted articles.

## Discussion

The most retracted articles belonged to the Diagnostic Pathology journal. All these 23 articles were retracted in 2016 which were published between 2013 and 2015 mainly because of concerns/issues about authorship, false/forged authorship, fake peer review, investigation by journal/publisher, misconduct by author, and plagiarism of text. The Tumor Biology journal approximately had the same results but the reasons for retraction differ. Biological Trace Element Research journal had the next rank with 13 retracted papers published between 2015-2019 which were retracted between 2017-2021 mostly because of concerns about data investigated by the company/institution. Iranian Red Crescent Medical Journal had 11 retracted articles in 2016 which were published in 2015 and 2016. The reasons for the retraction of articles from this journal were criminal proceeding for, false/forged authorship, fake peer review, the investigation by company/institution, investigation by journal/publisher, misconduct - official investigation/finding, and misconduct by author.

Mansourzadeh et al. (5) reviewed Iranian retracted publications published in PubMed up to December 2017. They discovered 164 articles which were mostly published in Diagnostic Pathology, Tumor Biology, and Iranian Red Crescent Medical Journal. They indicated that the mean lag time between publication and retraction was 20.8 months, ranging from less than a month to 8.4 years. These articles were mostly retracted because of these reasons; Authorship issues, plagiarism, redundant publication, no reason reported, overlap, misconduct, and honest error. In that study, Islamic Azad University and Tehran University of Medical Sciences had the highest number of retracted publications with 31 and 25 articles, respectively. Among these retracted articles, 142 publications were indexed in Web of Science and 161 in Scopus. They performed a citation analysis on 68 retracted publications indexed in Scopus between 2001 and 2013. These publications have received 789 citations (Citation per publication=11.6). Interestingly, 259 citations were identified after the retraction notice.

Masoomi et al. (6) in a study in 2018 estimated the percentage of retracted articles in Iran to be 0.1%. Although this percentage has increased compared to previous years, it is still a small percentage. But considering that these errors in publication can lead to wrong ethical judgments from organizations and countries, we suggest that more attention should be paid to reduce the retraction rate.

Ghorbi et al. (2) analyzed 343 Scopus-indexed articles which were retracted between 2001-2019. In their analysis, most articles were retracted from 2010 to 2016. The average time lag was 591 days after publication. Tumor Biology and Diagnostic Pathology had the highest number of retractions both of which are open-access journals. This study indicated that these articles were mostly retracted because of fake peer reviews, plagiarism, and duplicate publication. They reported that Islamic Azad University, University of Tehran, and Baqiyatallah University of Medical Sciences had the highest number of retractions. It seems that the second-highest retractions belong to Islamic Azad University (7).

## Conclusions

Publication of erroneous material and their subsequent retraction may result in important negative consequences including the loss of trust in scientific publications, induction of flawed information into further analyses and reviews, and misleading the academic community, among other problems. Due to the information above, ethics should be considered a more important issue in the process of conducting research projects and drafting the resultant articles. The fact that a noticeable number of articles that are written and even cited will be retracted shows a need for stronger filtration before their publication. We suggest that researchers, academic institutions, and journal editors and reviewers set higher standards while conducting research and clinical studies.

## Data Availability

All data produced in the present work are contained in the manuscript

## List of abbreviations

(COPE): Committee On Publication Ethics

## Declarations

### Consent for publication

Not applicable

### Availability of data and materials

The datasets used and/or analyzed during the current study are available from the corresponding author on reasonable request.

### Author Contributions

ShS, MN and AM search the database and collected the data. ShS, MN and AM wrote the manuscript. PS helped with writing the manuscript and scientific editing. All authors read and approved the final manuscript.

## Acknowledgments

This study was supported by the Isfahan University of Medical Sciences, Iran.

